# Clinical evaluation of BD Veritor^™^ SARS-CoV-2 point-of-care test performance compared to PCR-based testing and versus the Sofia^®^ 2 SARS Antigen point-of-care test

**DOI:** 10.1101/2020.09.01.20185777

**Authors:** Stephen Young, Stephanie N. Taylor, Catherine L. Cammarata, Celine Roger-Dalbert, Amanda Montano, Christen Griego-Fullbright, Cameron Burgard, Catherine Fernandez, Karen Eckert, Jeffrey C. Andrews, Huimiao Ren, Joseph Allen, Ronald Ackerman, Charles K. Cooper

## Abstract

**Objectives:** The clinical performance of the BD Veritor™ System for Rapid Detection of SARS-CoV-2 antigen (Veritor), a chromatographic immunoassay that detects the SARS-CoV-2 nucleocapsid antigen as a point-of-care test, was evaluated on nasal specimens from individuals with COVID-19 symptoms.

**Methods and Materials:** Two studies were completed to determine clinical performance. In the first study, nasal specimens and either nasopharyngeal or oropharyngeal specimens from 251 participants with COVID-19 symptoms (≤7 days from symptom onset [DSO]), ≥18 years of age, were utilized to compare Veritor with the Lyra^®^ SARS-CoV-2 PCR Assay (Lyra). In the second study, nasal specimens from 361 participants with COVID-19 symptoms (≤5 DSO), ≥18 years of age, were utilized to compare performance of Veritor to that of the Sofia^®^ 2 SARS Antigen FIA test (Sofia 2). Positive, negative, and overall percent agreement (PPA, NPA, and OPA, respectively) were the primary outcomes.

**Results:** In study 1, PPA for Veritor, compared to Lyra, ranged from 81.8%-87.5% for 0-1 through 0-6 DSO ranges. In study 2, Veritor had a PPA, NPA, and OPA of 97.4%, 98.1%, and 98.1%, respectively, with Sofia 2. Discordant analysis showed one Lyra positive missed by Veritor and five Lyra positives missed by Sofia 2; one Veritor positive result was negative by Lyra.

**Conclusions:** Veritor met FDA-EUA acceptance criteria for SARS-CoV-2 antigen testing (≥80% PPA point estimate) for the 0-5 and 0-6 DSO ranges. Veritor and Sofia 2 showed a high degree of agreement for SARS-CoV-2 detection. The Veritor test should facilitate rapid and reliable results for COVID-19 diagnosis utilizing easy-to-collect nasal swabs.

**Summary:** The BD Veritor SARS-CoV-2 antigen test met FDA-EUA acceptance criteria for SARS-CoV-2 antigen testing for subjects with COVID-19 symptoms (0-5 days post-onset). BD Veritor and Quidel Sofia 2 antigen tests had good agreement for SARS-CoV-2 detection; discordant analysis favored Veritor.

## INTRODUCTION

In response to the COVID-19 pandemic, an emphasis has been placed on SARS-CoV-2 diagnostic testing for symptomatic individuals.[1] Although laboratory-based PCR testing is considered the clinical reference standard for COVID-19 diagnosis, it is associated with some drawbacks, including false-negative reporting.[2-4] Also, limitations in capacity have been documented for PCR-based testing,[5, 6] which can lead to prolonged time to result (at best 24 hours when sample shipment is considered); and in most cases, dedicated staff and automated platforms are required to provide effective turn-around-time and optimized patient management.[7]

In February 2020, the World Health Organization identified point-of-care (POC) testing as a number one priority to address the COVID-19 pandemic.[8] Importantly, recent work has demonstrated that delays in test reporting can negatively impact the value of isolation as a control measure to reduce the spread of SARS-CoV-2.[9] The relatively small investment in resources and expertise required to perform POC testing makes it ideal for use in decentralized health care settings.[7] Antigen-based immunoassay POC tests for SARS-CoV-2 can target multiple viral antigens, including spike or nucleocapsid protein in a cartridge-based, lateral flow format. Although it is too early to determine whether one target is advantageous over another, evidence supports the efficacy of nucleocapsid detection in these types of antigen-based assays.[10, 11] Reports involving SARS and SARS-CoV-2 have demonstrated that the nucleocapsid protein is produced at high levels relative to the other viral proteins.[12, 13] In addition, nucleocapsid detection was recently shown, albeit in a serology-based test, to result in higher sensitivity for detection of SARS-CoV-2 compared to spike protein detection.[14]

US-FDA Emergency Use Authorization (EUA) was recently granted for the BD Veritor™ System for Rapid Detection of SARS-CoV-2 (henceforth referred to as “Veritor test”), a POC, chromatographic immunoassay that detects the SARS-CoV-2 nucleocapsid antigen. This report presents the performance data for the Veritor test using nasal swab specimens from COVID-19 symptomatic individuals compared to the Lyra**^®^** SARS-CoV-2 Assay (henceforth referred to as “Lyra assay”), which was utilized as the clinical reference standard. In a sub-population, Veritor test results were compared with results from another FDA-EUA authorized nucleoprotein antigen test, the Quidel Sofia^®^ 2 SARS Antigen FIA test (henceforth referred to as “Sofia 2 test”).

## METHODS AND MATERIALS

### Study design

Both studies described here involved a prospective collection of upper respiratory specimens. Eligible participants were ≥18 years of age and presented with one or more self-reported COVID-19 signs or symptoms. Individuals were excluded if a nasal swab was collected as part of standard of care (SOC). Demographic and healthcare-related information was collected (e.g. symptomology, health history, etc.). No study procedures were performed without an informed consent process or signature of a consent form. This research was performed in accordance with Good Clinical Practice guidelines and the Declaration of Helsinki. This article was prepared according to STARD guidelines for diagnostic accuracy studies reporting.[15]

### Specimen collection

#### Study 1 (EUA Veritor/Lyra comparison)

The first study was utilized to determine whether the Veritor test met FDA-EUA criteria for detection of SARS-CoV-2 in COVID-19 symptomatic individuals (within ≤7 DSO). Collection of specimens from 260 participants occurred across 21 geographically diverse study sites between June 5-11, 2020. Specimens for the Veritor test were from clinician-collected nasal specimens using regular-tipped flocked swabs (Becton, Dickinson and Company, BD Life Sciences—Integrated Diagnostics Solutions, Sparks, MD, USA) inserted approximately 2.5 cm up the nostril (from the edge of the nostril). The swab was rolled five times along the mucosa of the nostril to ensure that sufficient mucus and cells were collected; the process was repeated in the other nostril using the same swab.

Lyra assay specimens came from nasopharyngeal (NP) or oropharyngeal (OP) swabs; SOC OP or NP swabs were taken before any study swabs. If an NP swab was collected as part of SOC, the participant had the option of having an OP study swab taken in lieu of a second NP swab. All NP (n=217) or OP (n=34) specimens were clinician-collected. Reference testing was performed at TriCore Reference Laboratories while the Veritor testing was performed internally at BD (San Diego, CA, USA).

#### Study 2 (Veritor/Sofia 2 comparison)

The second study involved comparison of Veritor test performance to the Sofia 2 test for SARS-CoV-2 detection run with Sofia 2 analyzer. Collection occurred from 377 participants with symptoms of COVID-19 (≤5 DSO) from five study sites in the USA. Specimen collection for Veritor testing was performed as described above. For Sofia 2 testing, clinician-collected nasal specimens occurred using methods and swabs described in the IFU (Puritan^®^ regular foam swabs [Puritan, Guilford, ME, USA]). The specimens were obtained from a single nostril (with the most visible secretion) using gentle rotation. In some cases, due to an update in the Sofia 2 instructions for use (IFU), participants were instructed to blow their nose prior to nasal swab specimen collection. NP swab specimen collection for the Lyra assay (only for Veritor/Sofia 2 discordant testing) was performed as described above. Testing for Veritor, Sofia 2, and discordant Lyra assay, was performed at TriCore Reference Laboratories.

### Test procedures

Swabs were shipped for testing on dry ice (−70°C); nasal swabs were shipped dry and OP/NP swabs were shipped in universal viral transport medium. All testing was conducted with all personnel blinded to all other test results.

The Veritor and Sofia 2 tests were performed according to the manufacturer’s IFU (Becton, Dickinson and Company, BD Life Sciences—Integrated Diagnostic Solutions, San Diego, CA [16] and Quidel Corporation, Athens, OH,[17] respectively). Swabs were removed from −70°C storage ≤5 hours prior to the time of testing. Swabs were placed at 2-8°C for ≥2 hours and then at room temperature for 10-30 minutes prior to testing.

For specimen extraction prior to Veritor or Sofia 2 testing, the swabs were added to each respective extraction buffer tubes and mixed for at least 15-30 seconds or 1 minute, respectively. The extraction buffer/specimen mixture from each test was then added to the sample well of the corresponding test cartridge to initiate the testing. After the assays proceeded for 15 minutes, the test cartridges were inserted into either the Veritor or Sofia 2 analyzer to obtain results.

The Lyra assay was performed according to the manufacturer’s IFU (Quidel Corporation. Athens, OH).[18] When using the NucliSENS^®^ easyMAG^®^ and the Applied Biosystem 7500 Fast Dx Real-Time PCR instrument, the Lyra assay reports cycle number in a manner that omits the first 10 cycles; here the cycle numbers for the Lyra assay are reported with the first 10 cycles included. The BD MAX™ real time SARS-CoV-2 PCR assay (henceforth termed “MAX assay”) was used for discordant testing on residual nasal swabs following Veritor and Lyra testing in study 1. The MAX assay was performed according to the manufacturer’s IFU (Becton, Dickinson and Company, BD Life Sciences—Integrated Diagnostic Solutions, Sparks, MD).[19]

### Data collection and statistical analyses

The primary outcome measures for this study were positive, negative, and overall percent agreement (PPA, NPA, and OPA, respectively) point estimates for the Veritor test compared to results from the Lyra assay in study 1 and for the Veritor test compared to the Sofia 2 test in study 2.

For study 1, the acceptance criteria was a point estimate of ≥80% PPA of the Veritor test when compared to the Lyra assay; clinical evaluation required contiguous enrolment to a minimum of 30 prospectively collected positive specimens as specified in the Antigen Template for Manufacturers (May 11, 2020) for EUA submissions to the US-FDA.[20] Based on an estimated 10% prevalence rate, it was necessary to enroll approximately 300 participants to achieve the required number of positives.

For study 1, positive predictive value, negative predictive value, and accuracy were also calculated as secondary outcomes.[21] Additionally, a 2-sample t-test (2-tailed) was used to compare means between Lyra assay positive Ct values on specimens matched to Veritor negative and positive test results for SARS-CoV-2 in study 1.

## RESULTS

### Study 1 (EUA study)

#### Participant reconciliation, demographics, and COVID-19 symptomology

The mean and median age of the participants (44.7 and 43 years, respectively) were close (Table S1). More than half (64.2%) of the participants were female. By race, the largest proportion of participants were White, followed by Black, and then Asian. Approximately 40% were Hispanic or Latino. Cough was the most-reported symptom from participants, followed by muscle pain, and then headache. While the drive-through/tent and outpatient clinic collection site categories represented approximately three-fourths of the collection sites, the research clinic category had the highest positivity rate (22.2%). The mean for DSO among the participants was 3.2 days (Table S1). From 260 participants, six participants/participant specimen sets were removed due to inclusion/exclusion criteria non-compliance, and three were removed due to invalid specimens/results. Thus, 251 evaluable nasal specimens (each paired with either OP or NP specimens) were included (Figure S1a).

#### Veritor test performance and discordant reconciliation

Performance values for the Veritor test are shown by DSO, for participants providing valid specimens (Table 1). The 0-5 DSO range was the shortest range tested to have a PPA value above 80% and include at least 30 reference positive results. The 0-6 DSO range also met PPA value acceptance criteria. The NPA for the Veritor test was 100% for the 0-1 to 0-5 DSO ranges; however, the NPA value for the 0-6 and 0-7 DSO ranges was 99.5% (95% CI: 97.4, 99.9) (Table 1). The area under the curve (AUC) values associated with Veritor test performance for the 0-1 through the 0-6 DSO ranges were >0.9; the AUC value for the 0-7 DSO range was 0.88 (Table 1 and Figure 1). Performance values for the Veritor test compared to the Lyra Assay were analyzed by number of symptoms, as reported by participants during sample collection. As shown in Table 2, PPA point estimates were higher for the Veritor test when stratified by ≥2 symptoms versus 1 symptom for both the 0-5 DSO range (88.0% and 66.7%, respectively) and the 0-6 DSO range (88.9% and 57.1%, respectively). In addition, stratification of Lyra Ct scores (for the 38 positive reference specimens represented in the entire 0-7 DSO range) by 1 versus ≥2 symptoms showed overlapping distributions that were offset, with the 1 symptom Ct score distribution shifted towards higher Ct values (Figure 2a). The mean Ct value for the 1 symptom group (25.56), although not statistically different (p=0.077) from the ≥2 symptom mean Ct value (22.10), showed a trend towards having a higher mean Ct by approximately 3 cycles, an order of magnitude (Figure 2b).

**Table 1.** Veritor test performance at one through seven DSO

| Performance<sup>a</sup> | 1 DSO | 2 DSO | 3 DSO | 4 DSO | 5 DSO<sup>b</sup> | 6 DSO | 7 DSO |
| --- | --- | --- | --- | --- | --- | --- | --- |
| PPA %, [95% CI] | 87.5 [52.9, 97.8] | 85.0 [64.0, 94.8] | 81.8 [61.5, 92.7] | 85.2 [67.5, 94.1] | 83.9 [67.4, 92.9] | 82.4 [66.5, 91.7] | 76.3 [60.8, 87.0] |
| NPA %, [95% CI] | 100 [88.6, 100] | 100 [95.1, 100] | 100 [97.1, 100] | 100 [97.7, 100] | 100 [98.1, 100] | 99.5 [97.4, 99.9] | 99.5 [97.4, 99.9] |
| OPA %, [95% CI] | 97.4 [86.5, 99.5] | 96.8 [91.1, 98.9] | 97.3 [93.3, 99.0] | 97.9 [94.7, 99.2] | 97.8 [94.9, 99.1] | 97.1 [94.2, 98.6] | 96.0 [92.8, 97.8] |
| AUC | 0.94 | 0.93 | 0.91 | 0.93 | 0.92 | 0.91 | 0.88 |
| True positives | 7 | 17 | 18 | 23 | 26 | 28 | 29 |
| False negatives | 1 | 3 | 4 | 4 | 5 | 6 | 9 |
| True negatives | 30 | 75 | 127 | 162 | 195 | 210 | 212 |
| False positives | 0 | 0 | 0 | 0 | 0 | 1 | 1 |
| Total | 38 | 95 | 149 | 189 | 226 | 245 | 251 |

**Abbreviations:** DSO, days from symptom onset; PPA, positive percent agreement; NPA, negative percent agreement; OPA, overall percent agreement; AUC, area under the curve
<sup>a</sup>Performance of Veritor test compared to the Lyra assay as reference
<sup>b</sup>The Veritor test is FDA-authorized for detection of SARS-CoV-2 only in individuals that are 0-5 DSO

**Figure 1.**
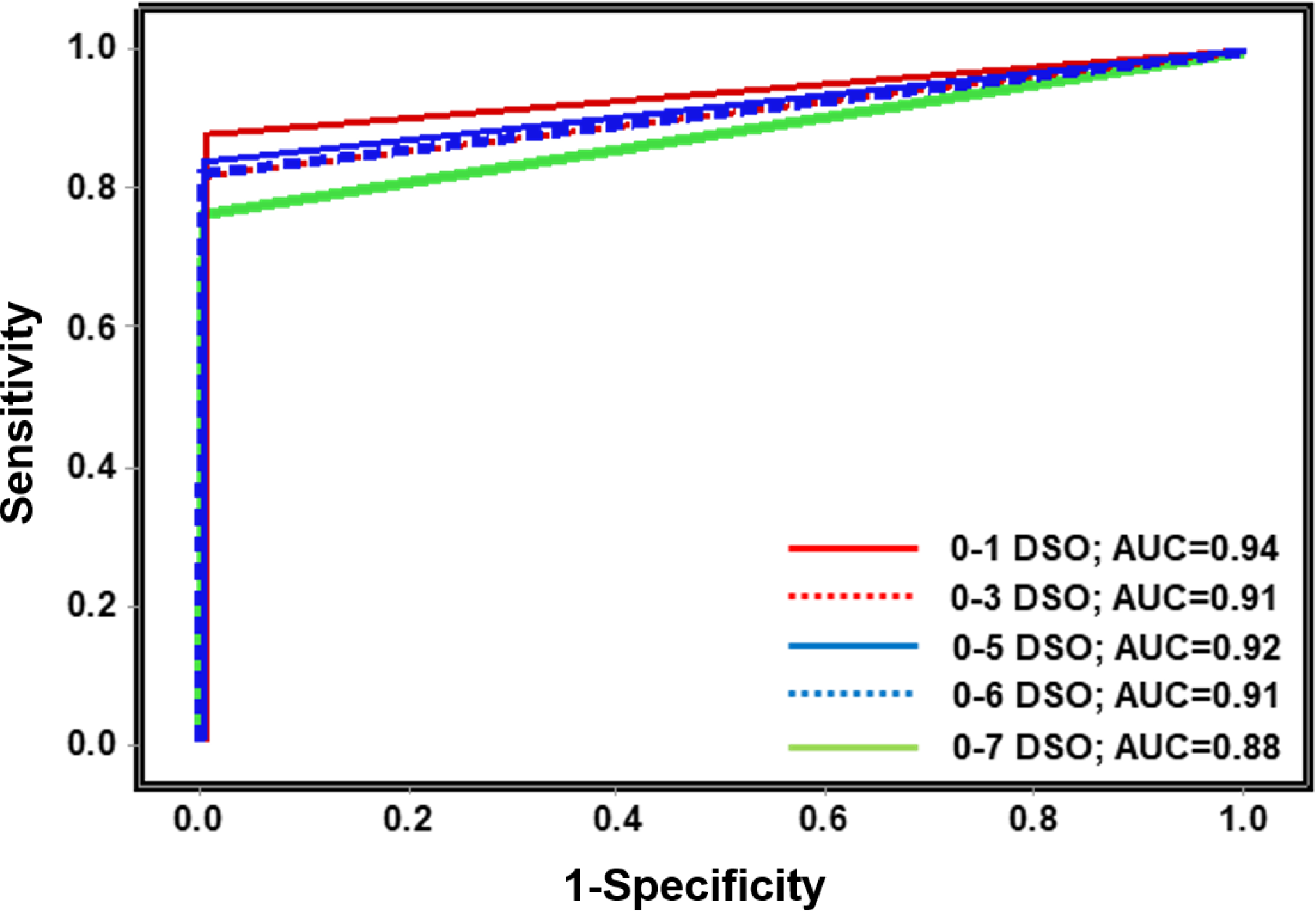
Veritor test performance results are plotted as a receiver-operator curve with sensitivity (corresponding to positive percent agreement) on the y-axis and 1-specificity (corresponding to 1-negative percent agreement) on the x-axis. Five lines, representing a 0-1 DSO, a 0-3 DSO, a 0-5 DSO, a 0-6 DSO, and a 0-7 DSO are shown. Also shown are the area under the curve values. **Abbreviations:** POC, point of care; DSO, days from symptom onset; AUC, area under the curve

**Table 2.** Veritor test performance by number of symptoms at 0-5 and 0-6 DSO

| Performance<sup>a</sup> | Number of symptoms |  |  |  |
| --- | --- | --- | --- | --- |
|  | 0-5 DSO |  | 0-6 DSO |  |
|  | 1 | ≥2 | 1 | ≥2 |
| PPA %, [95% CI] | 66.7 [30.0, 90.3] | 88.0 [70.0, 95.8] | 57.1 [25.0, 84.2] | 88.9 [71.9, 96.1] |
| NPA %, [95% CI] | 100 [95.7, 100] | 100 [96.6, 100] | 100 [95.8, 100] | 99.2 [95.6, 99.9] |
| OPA %, [95% CI] | 97.8 [92.3, 99.4] | 97.8 [93.7, 99.2] | 96.8 [91.0, 98.9] | 97.4 [93.4, 99.0] |
| True positives | 4 | 22 | 4 | 24 |
| False negatives | 2 | 3 | 3 | 3 |
| True negatives | 85 | 110 | 87 | 123 |
| False positives | 0 | 0 | 0 | 1 |
| Total | 91 | 135 | 94 | 151 |

**Abbreviations:** DSO, days from symptom onset; PPA, positive percent agreement; NPA, negative percent agreement; OPA, overall percent agreement
<sup>a</sup>Performance of Veritor test compared to the Lyra assay as reference

**Figure 2.**
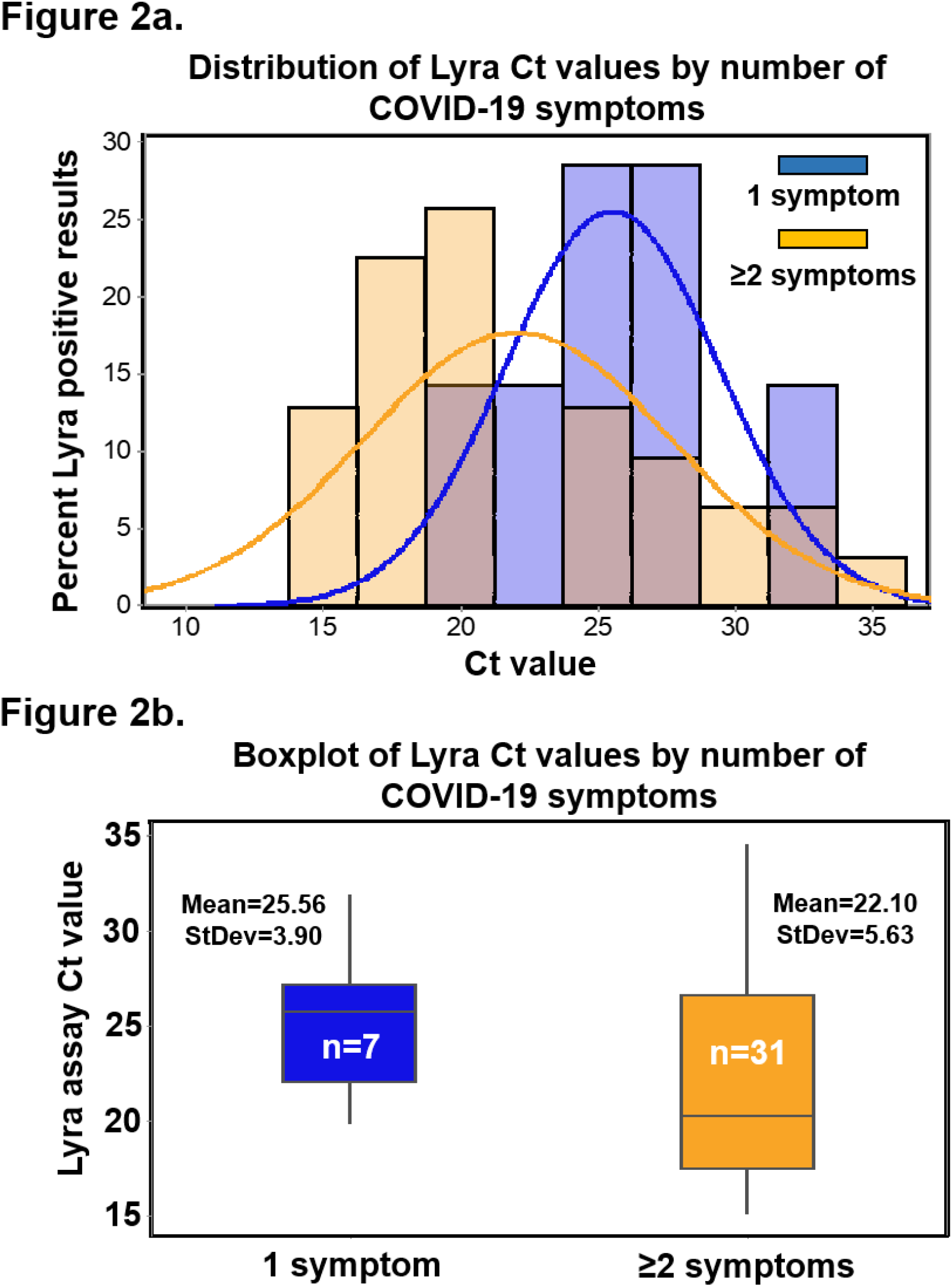
(a) The distribution of Ct values corresponding to the 38 specimens that were positive by the Lyra assay (from specimens collected from participants, 0-7 DSO) following stratification by number of symptoms. Ct score distribution for specimens matched to 1 symptom is shown in blue while those matched to ≥2 symptoms are shown in orange; the pink color indicates blue/orange overlap (b) The mean Ct values (and standard deviation) are shown for the ≥2 symptom specimens (n=31; mean=22.10, standard deviation=5.63) and the 1 symptom specimens (n=7; mean=25.56, standard deviation=3.90). A two-sample t-test (2-tailed) analysis indicated non-significant difference between the means (p-value=0.077; mean difference of 3.46; [95% CI: −0.43, 7.36]).

Eight of the nine false negative specimens by the Veritor test were from participants that had Lyra assay Ct values, which were greater than the mean Lyra Ct value (22.74); the ninth fell just below the mean value (Ct score of 22.04) (Figure 3a). The mean Ct value for Lyra assay results matched with the 29 true positive Veritor test results (20.76; standard deviation of 4.21) was significantly lower than the Lyra assay mean Ct value matched with the nine Veritor test negative discordant results (29.12; standard deviation of 4.11) (mean difference of 8.36; p-value <0.001; 2 sample t-test (2-tailed); 95% CI: 4.95, 11.77) (Figure 3b). MAX PCR assay testing showed a positive result for only two of the nine Veritor test negative discordant results (Table 3). From the remaining seven discordants, six were associated with a negative MAX assay result and one was associated with an unresolved result (no detection of internal control in the MAX assay).

**Figure 3.**
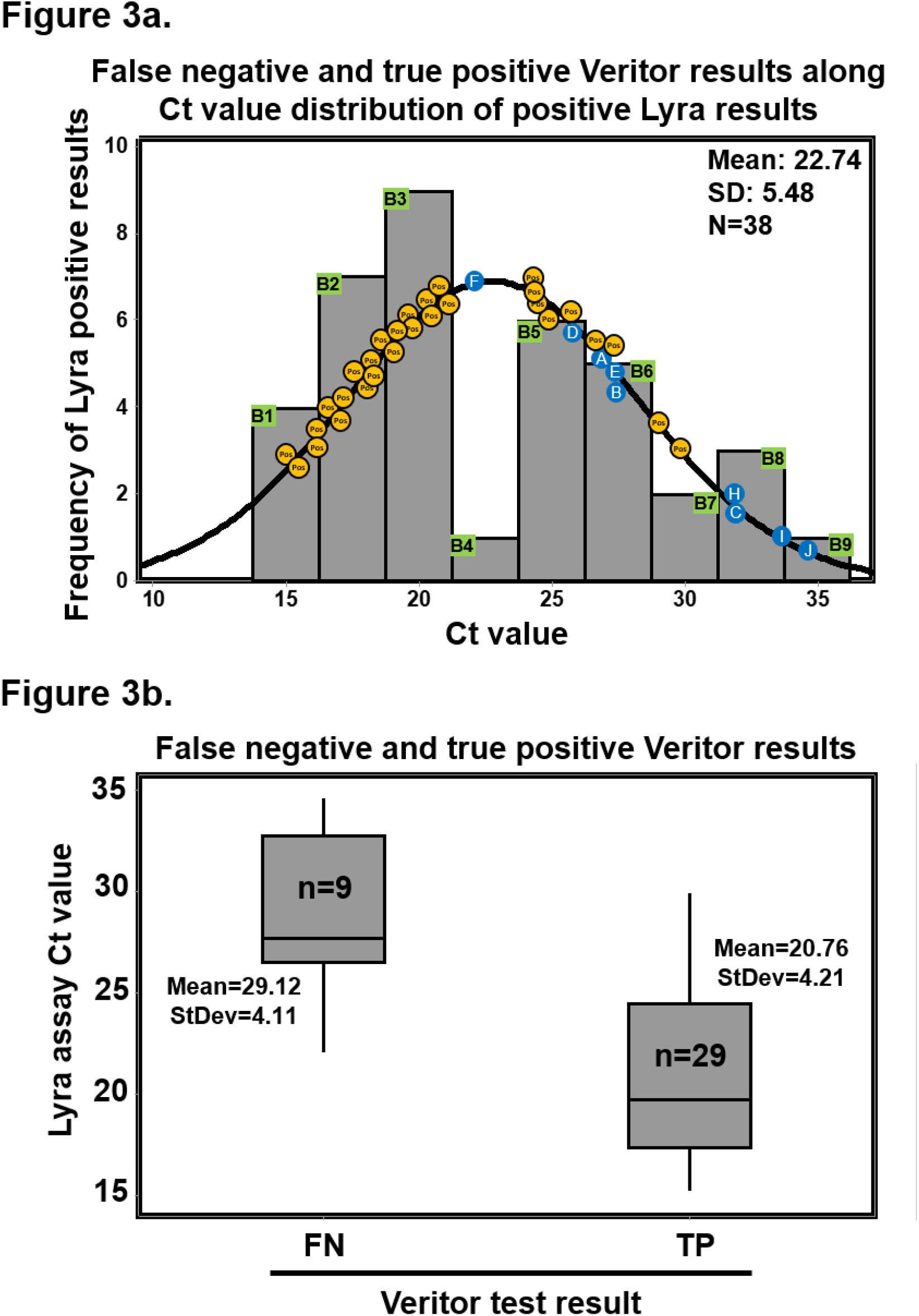
(a) The distribution of Ct values corresponding to the 38 specimens that were positive by the Lyra assay (from specimens collected from participants, 0-7 DSO). Plotted along the fitted distribution line are the 29 true positive Veritor results (orange circles) and the nine participant designations (letters superimposed onto blue circles), corresponding to those in Table 3, that represent the Veritor false negative results matched to Lyra assay Ct value. (b) The mean Ct values (and standard deviation) are shown for the 29 true positive (20.76 and 4.21, respectively) and the 9 false negative (29.12 and 4.11, respectively) Veritor test results. A two-sample t-test (2-tailed) analysis indicated a significantly higher mean Lyra assay Ct value for specimens matched to the 9 Veritor test false negative results compared to those matched to the 29 true positive results (p<0.001; mean difference of 8.36; [95% CI: 4.95, 11.77]).

**Table 3.** Discordant analysis for specimens associated with disagreement between Veritor test and Lyra assay

| DSO | Participant | Total FN | Total FP | Lyra result (Ct value) | Veritor result | MAX result (Ct value) | Serology result<sup>a</sup> |
| --- | --- | --- | --- | --- | --- | --- | --- |
| 0-1 | A | 1 | 0 | POS (27.21) | NEG<sup>b</sup> | NEG | n/a |
| 0-2 | B | 2 | 0 | POS (27.60) | NEG<sup>b</sup> | NEG | n/a |
|  | C | 3 | 0 | POS (31.90) | NEG<sup>b</sup> | NEG | n/a |
| 0-3 | D | 4 | 0 | POS (25.72) | NEG | POS (34.02) | POS: IgM and IgG |
| 0-4 | n/a | 4 | 0 | n/a | n/a | n/a | n/a |
| 0-5 | E | 5 | 0 | POS (27.56) | NEG<sup>b</sup> | NEG | n/a |
| 0-6 | F | 6 | 0 | POS (22.04) | NEG | UNR<sup>c</sup> | n/a |
|  | G | 6 | 1 | NEG (n/a) | POS | NEG | n/a |
| 0-7 | H | 7 | 1 | POS (31.84) | NEG | POS (32.72) | POS: IgM and IgG |
|  | I | 8 | 1 | POS (33.57) | NEG<sup>b</sup> | NEG | POS: IgM and IgG |
|  | J | 9 | 1 | POS (34.60) | NEG<sup>b</sup> | NEG | n/a |

**Abbreviations:** DSO, days from symptom onset; FN, false negative; FP, false positive; POS, positive; NEG, negative; UNR, unresolved
<sup>a</sup>Indicates serology testing done as part of standard of care prior to study-related activities
<sup>b</sup>Indicates agreement of Veritor test with MAX assay for a negative result for SARS-CoV-2
<sup>c</sup>Indicates a negative RNaseP result (internal control) in the MAX assay suggesting no presence of human material on the nasal swab

Figure 4 shows the PPV, NPV, and accuracy associated with the Veritor test by DSO range. As shown, PPV values for the Veritor test were 100%, for the 0-1 through 0-5 DSO ranges. There was only a single Veritor test positive/Lyra assay negative discordant result in the study, which occurred in the 0-6 DSO group and resulted in PPV point estimates of 96.6% and 96.7% for the 0-6 and 0-7 DSO ranges, respectively. The NPV values for the 0-1 to 0-6 DSO groups ranged from 96.8 to 97.2. At 0-7 DSO, the NPV was 95.9.

**Figure 4.**
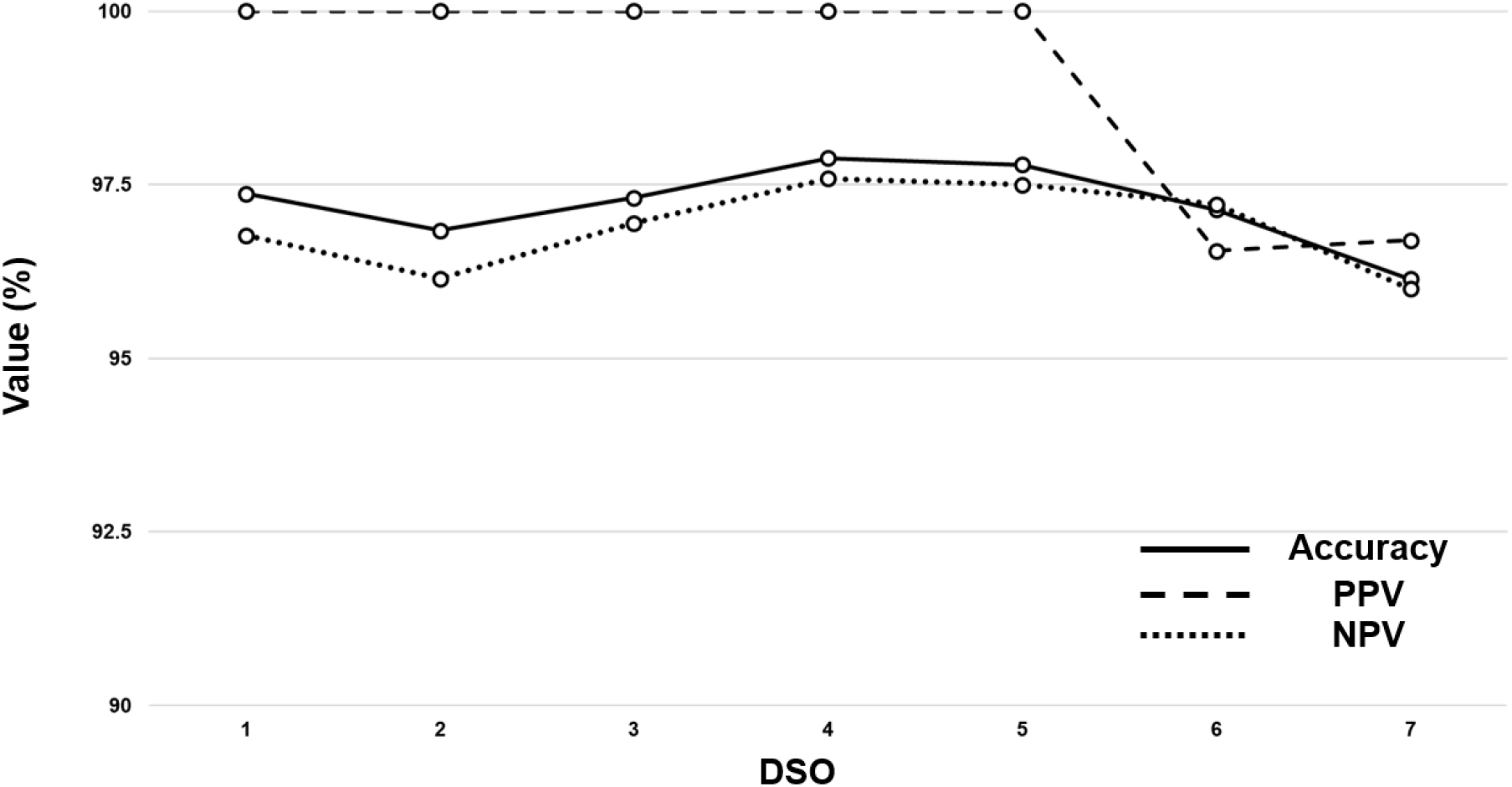
PPV, NPV, and test accuracy as a function of DSO for SARS-CoV-2 detection by the Veritor test. The point estimates for the three test values are plotted along the y-axis as percentages and the seven DSO ranges (0-1 to 0-7) reside along the x-axis. The SARS-CoV-2 prevalence value at each, respective, DSO range (based on positive reference results) was utilized for calculations and are as follows: 0-1 (21.1%), 0-2 (21.1%), 0-3 (14.8%), 0-4 (14.3%), 0-5 (13.7%), 0-6 (13.9%), and 0-7 (15.1%). Abbreviations: PPV, positive predictive value; NPV, negative predictive value; DSO, days from symptom onset.

### Study 2 (Veritor/Sofia 2 test comparison study)

#### Participant reconciliation, demographics, and COVID-19 symptomology

From 377 participants, four specimen sets were removed due to noncompliance with either inclusion or exclusion criteria, 16 were removed due to non-compliant specimens, specimen handling or transport, or invalid test results. There were 361 evaluable specimens included in analysis for this study (Figure S1b). The mean and median age of the participants (45.3 and 44 years, respectively) were similar. Fever, cough, headache, sore throat, and shortness of breath were the five most common symptoms reported (Table S2).

#### Veritor test performance and discordant reconciliation

The PPA, NPA, and OPA for the Veritor test compared to the Sofia 2 test using specimens at the 0-5 DSO range were 97.4 (95% CI: 86.5, 99.5), 98.1 (95% CI: 96.0, 99.1), and 98.1 (95% CI: 96.1, 99.1) (Table 4). Of the seven discordant results, one was Veritor negative/Sofia 2 positive and was positive by the Lyra assay; six were Veritor positive/Sofia 2 negative, with 5 being positive by the Lyra assay and one being negative by the Lyra assay.

**Table 4.**
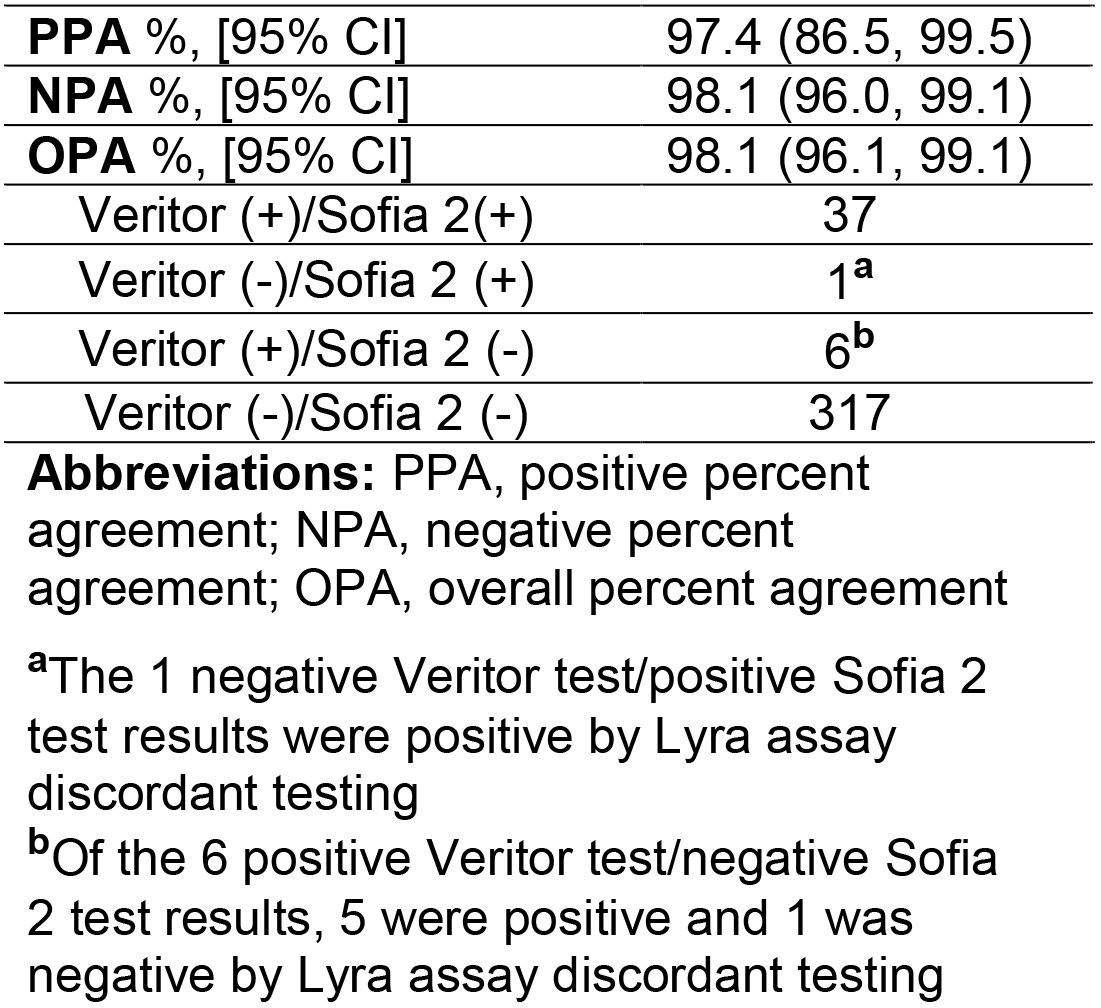
Agreement between Veritor and Sofia 2 for detection of SARS-CoV-2

|  |  |
| --- | --- |
| **PPA** %, [95% CI] | 97.4 (86.5, 99.5) |
| **NPA** %, [95% CI] | 98.1 (96.0, 99.1) |
| **OPA** %, [95% CI] | 98.1 (96.1, 99.1) |
| Veritor (+)/Sofia 2(+) | 37 |
| Veritor (-)/Sofia 2 (+) | 1<sup>a</sup> |
| Veritor (+)/Sofia 2 (-) | 6<sup>b</sup> |
| Veritor (-)/Sofia 2 (-) | 317 |

**Abbreviations:** PPA, positive percent agreement; NPA, negative percent agreement; OPA, overall percent agreement
<sup>a</sup>The 1 negative Veritor test/positive Sofia 2 test results were positive by Lyra assay discordant testing
<sup>b</sup>Of the 6 positive Veritor test/negative Sofia 2 test results, 5 were positive and 1 was negative by Lyra assay discordant testing

## DISCUSSION

The Veritor test was required to achieve ≥80% PPA relative to the reference standard (with at least 30 positive specimens by reference) in order to be considered acceptable for FDA-EUA. The Veritor test showed 83.9% and 82.4% PPA for specimens from COVID-19 symptomatic participants that were 0-5 and 0-6 DSO, respectively. In addition, the AUC values for the 0-1 through the 0-6 DSO ranges were excellent (ranging from 0.91-0.94). The results presented here suggest that the Veritor test should be effective in settings that would benefit from POC testing (e.g. decentralized health care settings) in order to classify 0-5 or 0-6 DSO individuals as positive or negative for SARS-CoV-2 infection to support patient management.

Discordant analysis for the 0-1 DSO through 0-6 DSO specimens revealed one false negative result (Participant D from Table 3) that was associated with a high (34.02) Ct value for the MAX assay. Interestingly, Participant D had a positive SOC serology result (both IgM and IgG), suggesting that the individual likely had a DSO greater than three. The nasal specimen from participant F had no detectable internal control (RNAse P gene), suggesting a lack of integrity for this specimen. The remaining four participants (A, B, C, and E) had nasal specimens that were negative by the MAX assay, agreeing with the Veritor test. The false-positive (participant G) Veritor test result had a line value that was close to the positive cutoff and was therefore a low positive.

Here the Veritor test had ≥96.0% PPV and NPV values for detection of the SARS-CoV-2 nucleocapsid antigen at all DSO ranges tested. Plotted values demonstrate the dependence of Veritor test NPV on disease prevalence (Table S3). Reflex testing (e.g. PCR-based testing) may be appropriate following a negative Veritor test result depending on the pretest probability and level of certainty required for patient management given medical history and future clinical action.

Discordant analysis for study 2 was performed using the Lyra assay and resulted in five Lyra and Veritor positive/Sofia 2 negative, one Lyra and Sofia 2 positive/Veritor negative, and one Veritor positive/Lyra and Sofia 2 negative result. For the latter result, the apparent false positive was associated with a Veritor test value that was close to the positive cutoff; this low positive was the lowest positive Veritor value observed in study 2.

PCR-based assays for diagnostic applications are typically highly sensitive for detecting target analyte relative to other diagnostic methods. However, recent results challenge whether this is always advantageous in all diagnostic settings. Bullard et al. (2020) and Wolfel et al. (2020) recently showed PCR-positive results at time points corresponding with negative culture-based testing for active SARS-CoV-2. Importantly, this discrepancy between testing methods seems to emerge around 6-8 DSO.[22, 23] In addition, Wolfel and colleagues show that the presence of sgRNA, a molecular marker for replicating SARS-CoV-2 virus, peaks around day 3-4 DSO, and then decreases drastically by day 6-7 DSO.[23] Finally, antigen-based test accuracy improves significantly when specimens associated with reference PCR values of 31-40 Ct are removed from analysis and only specimens matched with reference values of ≤30 Ct are included.[11] Eight of the nine false-negative Veritor test results here were matched with Lyra assay Ct values that were above the mean Ct value for the 38 Lyra assay positive results (four were approximately ten cycles above). This, combined with the significant difference in Lyra-matched Ct values for the 29 Veritor test true positive and 9 Veritor test false negative specimens, suggests that Veritor-to-Lyra concordance is indirectly proportional to the Lyra assay Ct score.

While PCR-based testing is sensitive for target detection, other testing modalities (such as antigen-based testing) may also be informative and may help clinicians determine the peak time frame during which infections are transmissible. However, more data is needed to establish the efficacy of antigen-based tests, such as Veritor or Sofia 2, for identifying contagious individuals—especially in the asymptomatic population. The Veritor and Sofia 2 tests are currently only authorized for individuals suspected of having a SARS-CoV-2 infection at 0-5 DSO. In addition, the high level of agreement observed between the Veritor and Sofia 2 tests as the tests is consistent with reported, similar limits of detection for SARS-CoV-2.[16, 17]

The difference in EUA labeled sensitivity for Sofia 2 (96.7%) vs Veritor (84%) was not supported by this study, probably due to spectrum differences in study design and patient populations in this study versus the Sofia 2 EUA study. The patient population chosen for this study was intended to reflect the performance of the Veritor test in clinical settings where decentralized POC testing such as antigen testing would be most appropriate. The study data presented here included a large proportion of specimens collected from clinical settings, such as drive-through testing, tents, and outpatient clinics, and therefore likely includes individuals with milder severity illness, compared with study populations that have been used to generate sensitivity estimates for other EUA antigen tests where enrollment included Emergency Department patients and hospitalized patients. Several publications have demonstrated an association between severe disease and higher viral loads, which could inflate antigen test sensitivity performance estimates when compared to performance estimates generated in patients with milder disease.[24-29] The finding in this study of an observed Ct score shift for subjects with 1 symptom vs ≥2 symptoms also supports the possibility that there may even be differences in viral load according to disease severity even amongst patients with milder disease. Analyses here (Table 2 and Figure 2) suggest that ≥2 symptoms also demonstrated a higher PPA than 1 symptom alone, which is reflective of the by a trend towards lower Ct scores (higher viral load) for specimens from participants with ≥2 symptoms.

### Limitations

This study had some limitations. First, the Veritor test was performed on nasal swab specimens; however, the Lyra assay was performed on either NP (or OP) swab specimens per FDA-EUA requirements. Other EUA submissions (the LumiraDx SARS-CoV-2 Ag Test [“Luminar test”] and the Abbott BinaxNOW COVID-19 Ag CARD [“Abbott test”]) utilized nasal swab specimens for both the antigen test and the reference PCR assay. Furthermore, MAX from the remnant Veritor nasal swab in this report agreed with negative Veritor results in 7 of 9 discordant specimens. Improved PPA for Veritor versus Lyra may have been achieved through the use of paired nasal swab specimens in the EUA study.

The Sofia 2 assay in study 2 was performed on nasal swabs that were collected either with (n=70; Table S4), or without (Table S5), a nose blowing step prior to collection. The nose-blowing step was an addition to the Sofia 2 test IFU intended only to reduce the frequency of invalid results (by reducing the amount of mucousal-, or blood-derived inhitors in the specimen), and was not included in order to alter the performance of the Sofia 2 test. Although the n is low for specimens with a pre-nose blowing step in study 2, here, the results suggest that the nose-blowing step did not alter the overall performance of the Sofia 2 test in relation to the Veritor test.

### Conclusions

The Veritor test met acceptance criteria for Emergency Use Authorization criteria for antigen testing (≥80% PPA point estimate) for the 0-5 and 0-6 DSO ranges in a population of 251 subjects. The 0-1 through 0-6 DSO ranges had AUC values ≥0.90, suggesting that it is a reliable point of care test. Results here suggest that number of symptoms may influence the sensitivity of antigen-based POC testing. In additional testing, Veritor returned 43 positive results and Sofia 2 returned 37 positive results from a population of 361 subjects. The speed (15 minute run time) and performance of antigen tests for SARS-CoV-2 detection should facilitate rapid and reliable results for COVID-19 diagnosis. Importantly, this POC test is run on nasal swab specimens, which are relatively easy and safe to collect. This study generated point estimates from a population that represents the most appropriate intended use population and thus can be used to inform proper patient management. In addition, the Veritor test should have a significant impact in decentralized healthcare settings where requirements for larger-scale PCR-based tests are harder to meet or result in extended turn-around-times.

## Data Availability

The data that support the findings of this study should be requested from Dr. Charles K. Cooper. Contact

## FUNDING

This study was funded by Becton, Dickinson and Company; BD Life Sciences—Integrated Diagnostics Solutions. Non-BD employee authors received research funds as part of this work.

## CONFLICTS OF INTEREST

The authors disclose the following conflicts of interest:

CRD, CF, KE, JCA, HR, and CKC are employees of Becton, Dickinson and Company; SY, None; CC, None; AM, None; CGF, None; CB, None; JA, None; RA, CEO and PI of Comprehensive Clinical Research LLC

## ACKNOWLEDGEMENTS

The authors would like to thank Richard Anderson, Dave Kurisko, Edith Torres-Chavolla, Katherine Christensen, and Devin S. Gary (Becton, Dickinson and Company, BD Life Sciences – Diagnostic Systems), for their input on the content of this manuscript and editorial assistance. The authors also thank Stanley Chao and Yongqiang Zhang (Becton, Dickinson and Company, Global Clinical Development – Statistics & Clinical Data) for statistical support. The individuals acknowledged here have no additional funding or additional compensation to disclose.

## Supplemental Material

**Table S1.** Study 1 (EUA) participant demographics (N=254)

| **Age, years** | **Value** |
| --- | --- |
| Mean | 45.0 |
| SD | 16.6 |
| Median | 43.0 |
| Min | 18 |
| Max | 90 |
| **Gender, % (n)** |  |
| Female | 64.2 (163) |
| Male | 35.8 (91) |
| **Race, % (n)** |  |
| Asian | 5.9 (15) |
| Black | 10.2 (26) |
| White | 56.7 (144) |
| Other | 27.2 (69) |
| **Ethnicity, % (n)** |  |
| Hispanic or Latino | 40.6 (103) |
| Not Hispanic or Latino | 55.9 (142) |
| Not reported | 3.5 (9) |
| **Symptoms, % (n)** |  |
| Cough | 43.3 (110) |
| Muscle pain | 38.6 (98) |
| Headache | 37.4 (95) |
| Sore throat | 35.4 (90) |
| Fever | 30.7 (78) |
| Shortness of breath | 20.5 (52) |
| Chills | 17.7 (45) |
| New loss of taste or smell | 4.7 (12) |
| Repeated shaking with chills | 4.3 (11) |
| Other<sup>a</sup> | 24.0 (61) |
| **Collection site** |  |
| **All Sites<sup>b, c</sup>** | 21 |
| **Drive through/tent** (% of total sites) | 23.8 (5/21) |
| Total specimens collected | 42 |
| % REF positive of total collected | 2.4 (1/47) |
| **Outpatient clinic** (% of total sites) | 52.4 (11/21) |
| Total specimens collected | 74 |
| % REF positive of total collected | 24.3 (18/74) |
| **Research clinic** (% of total sites) | 19.0 (4/21) |
| Total specimens collected | 72 |
| % REF positive of total collected | 22.5 (16/71) |
| **Skilled nursing facility** (% of total sites) | 4.8 (1/21) |
| Total specimens collected | 66 |
| % REF positive of total collected | 4.5 (3/66) |
| **Overall positivity rate** | 15.1 (38/252) |
| **Mean DSO** | 3.2 |
| **Median DSO** | 3.0 |
| **Mode DSO** | 2.0 |

**Abbreviations:** SD, standard deviation; MIN, minimum; Max, maximum; DSO, days from symptom onset; REF, reference test
<sup>a</sup>"Other" symptoms include gastrointestinal issues, fatigue, chest pain, rhinorrhea, and nasal congestion
<sup>b</sup>Collection sites reside in the following states: Arizona, California, Florida (7), Georgia, Iowa, Louisiana, North Carolina, Nevada (2), Ohio (3), South Carolina, Texas, Utah
<sup>c</sup>% REF positive of total collected values were calculated from specimens only with valid Lyra assay results (n=252)

**Table S2.** Study 2 (Veritor vs Sofia 2 comparison) participant demographics (N=373)

| Age, years | Value |
| --- | --- |
| Mean | 45.4 |
| SD | 16.4 |
| Median | 44 |
| Min | 18 |
| Max | 98 |
| **Gender, % (n)** |  |
| Female | 59.5 (222) |
| Male | 40.5 (151) |
| **Race, % (n)** |  |
| Asian | 0.3 (1) |
| Black | 16.1 (60) |
| White | 69.4 (259) |
| Other | 14.2 (53) |
| **Ethnicity, % (n)** |  |
| Hispanic or Latino | 66.2 (247) |
| Not Hispanic or Latino | 33.8 (126) |
| **Symptoms, % (n)** |  |
| Cough | 56.3 (210) |
| Muscle pain | 53.9 (201) |
| Headache | 67.8 (253) |
| Sore throat | 52.8 (197) |
| Fever | 34.3 (128) |
| Shortness of breath | 13.7 (51) |
| Chills | 37.5 (140) |
| New loss of taste or smell | 17.7 (66) |
| Repeated shaking with chills | 1.6 (6) |
| Other<sup>a</sup> | 43.7 (163) |
| **Collection site** |  |
| **All Sites** | 5 |
| **Drive through/tent (% of total sites)** | 20.0 (1/5) |
| Veritor positivity rate (%) | 11.1 (1/9) |
| Sofia 2 positivity rate (%) | 11.1 (1/9) |
| **Outpatient clinic (% of total sites)** | 20.0 (1/5) |
| Veritor positivity rate (%) | 8.2 (4/49) |
| Sofia 2 positivity rate (%) | 6.1 (3/49) |
| **Research clinic (% of total sites)** | 40.0 (2/5) |
| Veritor positivity rate (%) | 12.8 (38/298) |
| Sofia 2 positivity rate (%) | 11.4 (34/297)<sup>b</sup> |
| **Urgent care (% of total sites)** | 20.0 (1/5) |
| Veritor positivity rate (%) | 0.0 (0/7) |
| Sofia 2 positivity rate (%) | 0.0 (0/7) |
| **Overall (either test) positivity rate** | 12.1 (44/364) |
| **Mean DSO** | 3.0 |
| **Median DSO** | 3.0 |
| **Mode DSO** | 4.0 |

**Abbreviations:** SD, standard deviation; MIN, minimum; Max, maximum; DSO, days from symptom onset; REF, reference test
<sup>a</sup>"Other" symptoms include shaking, gastrointestinal issues, fatigue, chest pain, rhinorrhea, and nasal congestion
<sup>b</sup>Of the 298 specimens, one was Sofia 2 invalid

**Table S3.**
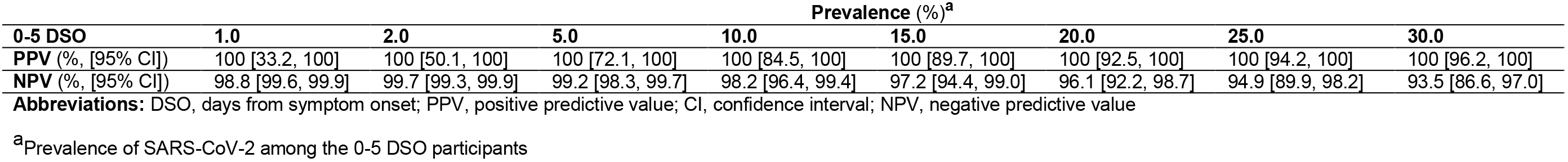
Positive and negative predictive values for the Veritor test compared to the Lyra Assay

**Table S4.** Agreement between Veritor and Sofia 2 (only collection involving nose blowing prior to nasal swab collection) for detection of SARS-CoV-2

|  |  |
| --- | --- |
| **PPA** %, [95% CI] | 100 (56.6, 100) |
| **NPA** %, [95% CI] | 98.0 (89.7, 99.7) |
| **OPA** %, [95% CI] | 98.2 (90.6, 99.7) |
| Veritor (+)/Sofia 2(+) | 5 |
| Veritor (-)/Sofia 2 (+) | 0 |
| Veritor (+)/Sofia 2 (-) | 1<sup>a</sup> |
| Veritor (-)/Sofia 2 (-) | 50 |

**Abbreviations:** PPA, positive percent agreement; NPA, negative percent agreement; OPA, overall percent agreement
<sup>a</sup>The 1 positive Veritor test/negative Sofia 2 test result was positive by Lyra assay discordant testing

**Table S5.**
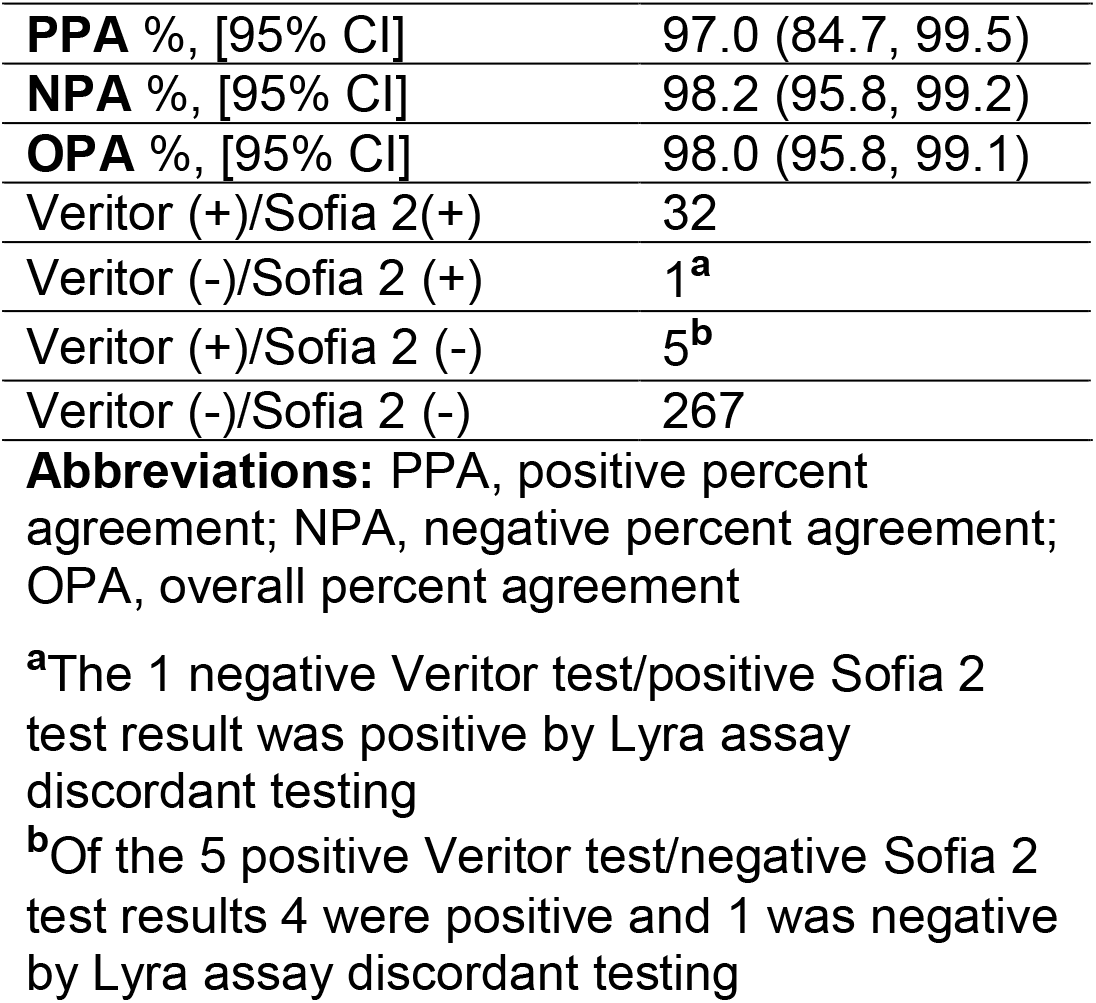
Agreement between Veritor and Sofia 2 (nose blowing not included prior to nasal swab collection) for detection of SARS-CoV-2

**Figure S1.**
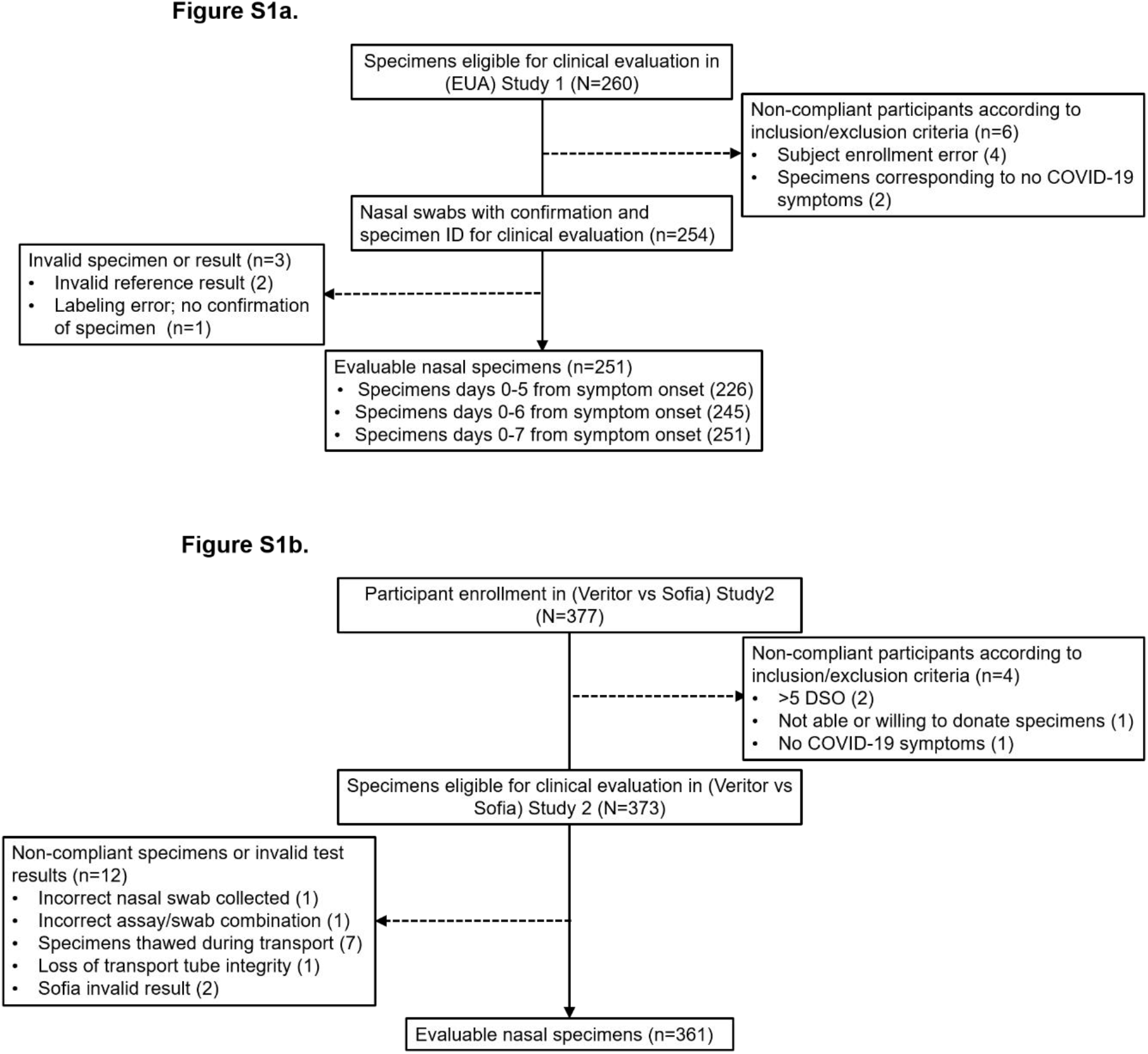
**(a)** Reconciliation during enrollment of swab specimens from participants, ≥18 years of age, with signs or symptoms of COVID-19 for study 1. **Abbreviations:** ID, identification; DSO, days from symptom onset **(b)** Reconciliation during enrollment of swab specimens from participants, ≥18 years of age, with signs or symptoms of COVID-19 for study 2. **Abbreviations:** DSO, days from symptom onset

## Notes

### Clinical Trial

Registration of this study at ClinicalTrials.gov was not obtained as it did not fulfill the applicable minimum medical device clinical trial requirement criteria as per the Food and Drug Administration Amendments Act. Specifically, this study did not assess health outcomes associated with the use of this assay.

### Author Declarations

Continuing review approval: Advarra IRB-CR00170042-Approval date: 3 Dec 2019 Continuing review Becton Dickinson - BDX-GSCP01, Becton Dickinson Program for Collection of BioSpecimens (Pro00015576) study 1 COVID approval notice: Advarra IRB-MOD00683499-Approval date: 29 May 2020 Becton Dickinson - BDX-GSCP01, Becton Dickinson Program for Collection of BioSpecimens (Pro00015576) Study 2 COVID approval notice: Advarra IRB-MOD00717838-Approval date: 13 Jul 2020 Becton Dickinson - BDX-GSCP01, Becton Dickinson Program for Collection of BioSpecimens (Pro00015576)

